# Detection and categorization of severe cardiac disorders based solely on pulse interval measurements

**DOI:** 10.1101/2022.01.10.22268961

**Authors:** Shigeru Shinomoto, Yasuhiro Tsubo, Yoshinori Marunaka

## Abstract

Cardiac disorders are common conditions associated with a high mortality rate. Due to their potential for causing serious symptoms, it is desirable to constantly monitor cardiac status using an accessible device such as a smartwatch. While electrocardiograms (ECGs) can make the detailed diagnosis of cardiac disorders, the examination is typically performed only once a year for each individual during health checkups, and it requires expert medical practitioners to make comprehensive judgments. Here we describe a newly developed automated system for alerting individuals about cardiac disorders solely based on pulse interval measurements. For this purpose, we examined two metrics of heart rate variability (HRV) and analyzed 1-day ECG recordings of more than 1,000 subjects in total. We found that a newly introduced metric of local variation was more efficient than conventional HRV metrics for detecting premature contraction, and furthermore, that a suitable combination of the old and new metrics resulted in much superior detectability particularly for atrial fibrillation, which requires more attention. Even with a 1-minute recording of pulse intervals, our new detection system had a diagnostic performance even better than that of the conventional analysis method applied to a 1-day recording.

## I. INTRODUCTION

Heart disease has one of the highest mortality rates of any condition, which is similar to that of cancer [1–5]. Early detection of cardiac disorders might help prevent fatalities. One of the most basic means of detecting irregularities is pulse assessment, which was first used in ancient China and Egypt in centuries B.C. [6–8]. In the modern era, the electrocardiogram (ECG) provides a detailed assessment of cardiac electrical activity, reflecting depolarization and repolarization of the atria and ventricles [9]. Based on this information, a variety of diagnoses can be made, such as premature ventricular contraction (PVC), premature atrial contraction (PAC), and atrial fibrillation (AF).

Though efficient, ECG examinations are costly and inconvenient. While electrical signals are efficiently analyzed by automated algorithms, comprehensive diagnoses must be made by medical practitioners in the end [10, 11]. Accordingly, they are typically carried out only once a year for each individual during physical checkups, and fewer people are recommended to perform Holter ECG examinations continuously for 1 day, only if some cardiac anomalies are found in the physical checkups. More serious is that those who were negative in those ECG examinations still have a chance of getting cardiac disorders.

Thus it is desirable to make it possible to alert all persons to cardiac dysfunction using a handy device such as a smartwatch. While full ECG information is not available, a handy device may make it possible to constantly monitor a series of pulse intervals, which may provide sufficient information for detecting cardiac disorders [12]. This has led us to an idea of identifying cardiac disorders by analyzing heart rate variability (HRV) through pulse intervals in as much detail, just as expert medical practitioners have been doing for centuries.

HRV is quantified by measuring fluctuations in heartbeat intervals, using metrics such as the standard deviation of intervals between successive cardiac R waves or RR intervals (SDRR) and the coefficient of variation (*Cv*), which is defined as the ratio of the standard deviation to the mean [13, 14]. Nevertheless, the variation in heartbeat intervals is caused by not only cardiac disorders but also slow fluctuations in the heart rate that occur even in healthy people [15, 16]. There have been many attempts for isolating cardiac disorders from such miscellaneous factors, using a variety of analysis methods such as linear, frequency domain, wavelet domain, and nonlinear methods [17–21]. Nevertheless, diagnosing cardiac symptoms still requires medical experts to consider various possible conditions when making comprehensive judgments.

Recently, artificial intelligence-aided methods based on deep learning algorithms have been adopted for detecting cardiac disorders from heartbeat signals [5, 22–25]. While these methods achieve high performance, individual decisions depend on a huge number of parameters that were empirically determined with given training datasets. A problem of such artificial intelligence-aided methods is that we cannot explain the reason for individual diagnoses made for each case. Another problem is that such machine-learning diagnoses might depend sensitively on given training datasets.

To improve these points, here we design an automatic diagnosing system by simply combining two metrics that measure specific aspects of heartbeat variability. On a plane of the two metrics, we specify decision boundaries for different types of cardiac disorders. From this, we can characterize the difference in various cardiac diseases explicitly in terms of two metrics.

As the first metric, we have adopted the metric of local variation (*Lv*), which was previously used to analyze neuronal firing in the brain [26–28]. This is because *Lv* captures the difference in adjacent pulse intervals by removing the influence of slow fluctuations in heart rate, the latter of which might have disturbed the conventional HRV analysis. We have confirmed that *Lv* was superior to the conventional *Cv* in detecting premature contraction. Nevertheless, we also realized that *Cv* may still have detected some aspect independent of *Lv* particularly for atrial fibrillation. By identifying the differences between *Lv* and *Cv*, we sought to combine them in a manner that would maximize the ability to characterize cardiac status. Through this process, we invented a new metric, the local-global variation ratio (*Lg*), that effectively discriminates between premature contractions and fibrillation. Using these two metrics *Lv* and *Lg*, we constructed an algorithm to automatically diagnose cardiac symptoms. The new method exhibits outstanding performance in detecting AF, which is associated with serious diseases [29–32].

For this analysis, we used Holter ECG recordings obtained from more than 1,000 outpatients in total who had cardiological examinations at Clinic, Kyoto Industrial Health Association. Recordings contain a series of RR intervals monitored for 1 day, and are accompanied by diagnoses made by medical doctors and technicians, including not only AF but also PVC and PAC, the latter of which may take place even in healthy people [33, 34]. By assuming that pulse intervals that can be measured by a handy device are noisy observation of RR intervals [35, 36], we attempted to infer diagnoses of PVC, PAC, and AF from a set of RR intervals and examined how the inference can withstand the possible fluctuations added to the RR intervals, mimicking the noisy observations. To examine the generality of the analysis, we also applied the current method to the publicly available MIT-BIH databases, which have been conventionally used as standard datasets.

## II. RESULTS

For Holter ECG data recorded from 1,017 subjects in total (863 independent persons), we computed the variability metrics from each series of RR intervals and compared their ability to detect cardiac conditions, such as PVC, PAC, and AF.

### A. Comparison of variability metrics

Figure 1 depicts the distributions of PVC, PAC, and AF plotted against the RR interval metrics: the heart rate *Hr*, the logarithm of coefficient of variation log_10_ *Cv*, and the logarithm of the local variation log_10_ *Lv* (METHODS), each of which is calculated every 10 minutes and averaged over 18 hours. Because *Cv* and *Lv* computed for heartbeats are much smaller than unity, we take their logarithm to focus on their difference.

**FIG. 1.**
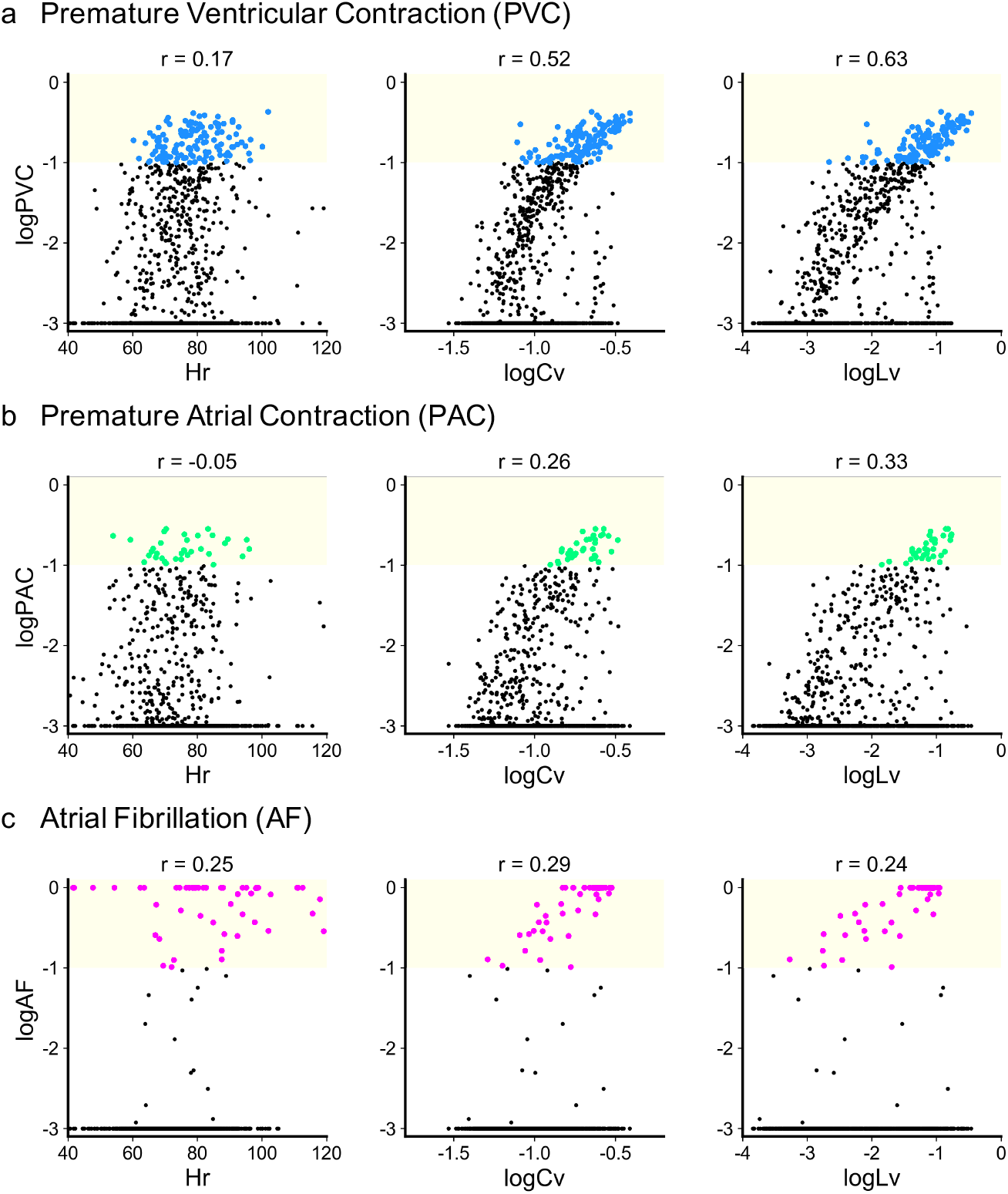
Cardiac disorders plotted against *Hr*, log_10_ *Cv*, and log_10_ *Lv*. Each dot represents the average of 10-minute statistics for a single subject over 18 hours. Vertical axes are the logarithm of (a) premature ventricular contraction (PVC); (b) premature atrial contraction (PAC); and (c) atrial fibrillation (AF). Values of the Pearson correlation *r* are indicated above. Yellow zones represent regions of *>* 0.1.

Many subjects had nonzero PVC and PAC, which were determined by an automatic detection algorithm; those with values higher than 10^−3^ comprised 49% and 33% of the entire subjects, respectively, and their levels were distributed rather continuously. In contrast with these, only 72 of 1,017 subjects (7%) exhibited nonzero AF over 18 hours, as determined manually by expert medical technicians.

For PVC, PAC, and AF, we categorized the status of each subject as 0 or 1 according to whether the ratio of each condition was lower or higher than a threshold of 0.1, and predicted the dichotomic status based on *Cv* or *Lv* as obtained from pulsation signals. The numbers of the 1,017 subjects at high risk of PVC, PAC, and AF were 128 (12%), 34 (3%), and 53 (5%), respectively. Only one subject (0.1%) exhibited multiple symptoms (PAC and AF).

A conventional HRV metric *Cv* and our newly introduced metric *Lv* were strongly correlated with cardiac symptoms; subjects who exhibited the higher *Cv* or *Lv* were more likely to exhibit cardiac disorders. The average heart rate *Hr* was relatively weakly correlated with cardiac symptoms, and accordingly, we shall concentrate on *Cv* and *Lv* in the following analysis.

We first examined whether the high value of *Cv* or *Lv* could signal each cardiac disorder. We categorized data as “positive” or “negative” according to whether or not log_10_ *Cv* or log_10_ *Lv* was greater than a given threshold. Positive data were categorized as “true positive (TP)” or “false positive (FP)” according to whether or not subjects exhibited a given cardiac disorder (PVC, PAC, or AF). In contrast, negative data were categorized as “false negative (FN)” or “true negative (TN)” according to whether or not subjects exhibited a given cardiac disorder.

The upper rows in Fig. 2 depicts the true positive ratio TPR = *N*_TP_*/*(*N*_TP_ + *N*_FN_), and the true negative ratio TNR = *N*_TN_*/*(*N*_TN_ + *N*_FP_), where *N*_TP_, *N*_FN_, *N*_TN_, and *N*_FP_ are the numbers of true-positive, false-negative, true-negative, and false-positive cases, respectively. Higher *Cv* or *Lv* results in higher TNR, implying that subjects in whom the metric is lower than the threshold are unlikely to exhibit cardiac disorders. However, a high threshold lowers TPR, implying that many cardiac disorders are missed.

**FIG. 2.**
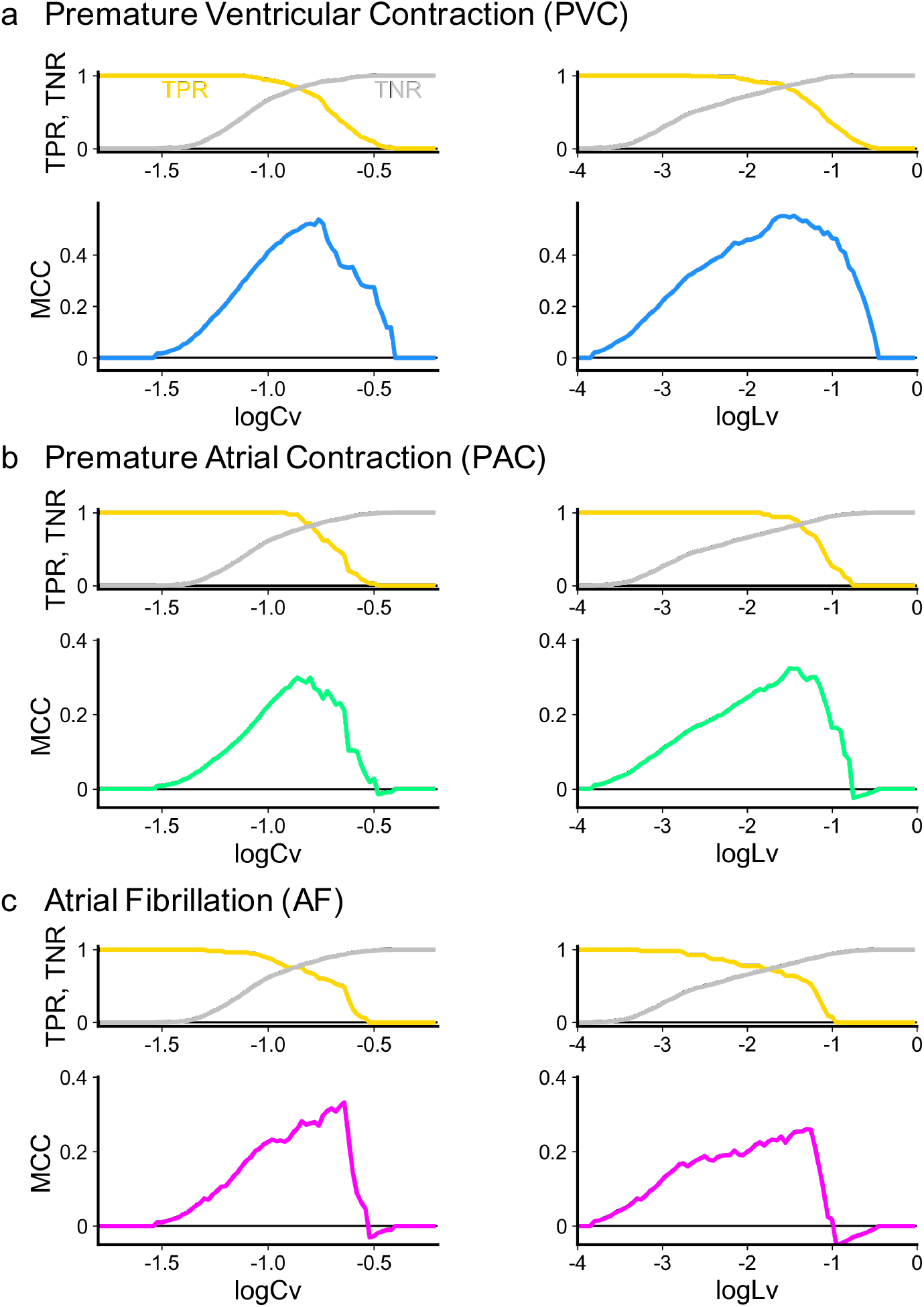
The true positive rate (TPR), true negative rate (TNR), and the Matthews correlation coefficient (MCC) measuring the ability of two individual metrics, thresholds of log_10_ *Cv* and log_10_ *Lv*, to detect the following cardiac disorders: (a) PVC, (b) PAC, and (c) AF.

One means of achieving a reasonable balance between true and false positives and negatives is to maximize the Matthews correlation coefficient (MCC) [37] defined as

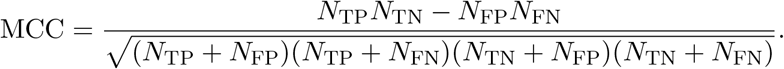

The lower rows in Fig. 2 depict the MCC values for PVC, PAC, and AF; peaks occur at an intermediate value for a threshold of log_10_ *Cv* or log_10_ *Lv*. For PVC and PAC, the new metric *Lv* is more efficient than *Cv* in achieving higher MCC values. By contrast, *Cv* results in a higher MCC than *Lv* for AF.

### B. Improving the detection of cardiac disorders

In the aforementioned analysis, we have seen that *Cv* and *Lv* had different strengths for different disorders. Because these metrics capture different aspects of the RR-variability, their performance in detecting cardiac disorders might be increased if these metrics were suitably combined.

Figure 3(a) displays the distribution of datapoints representing subjects with different cardiac diseases plotted on a plane spanned by log_10_ *Lv* and log_10_ *Cv*. The distribution seemingly comprises different groups; there is one big cluster at the lower left, centered at low log_10_ *Lv* ≈ −3 and low log_10_ *Cv* ≈ −1.2, while the datasets with higher log_10_ *Lv >* −2 tended to align linearly with the slope in parallel to log_10_ *Lv* − 2 log_10_ *Cv*.

**FIG. 3.**
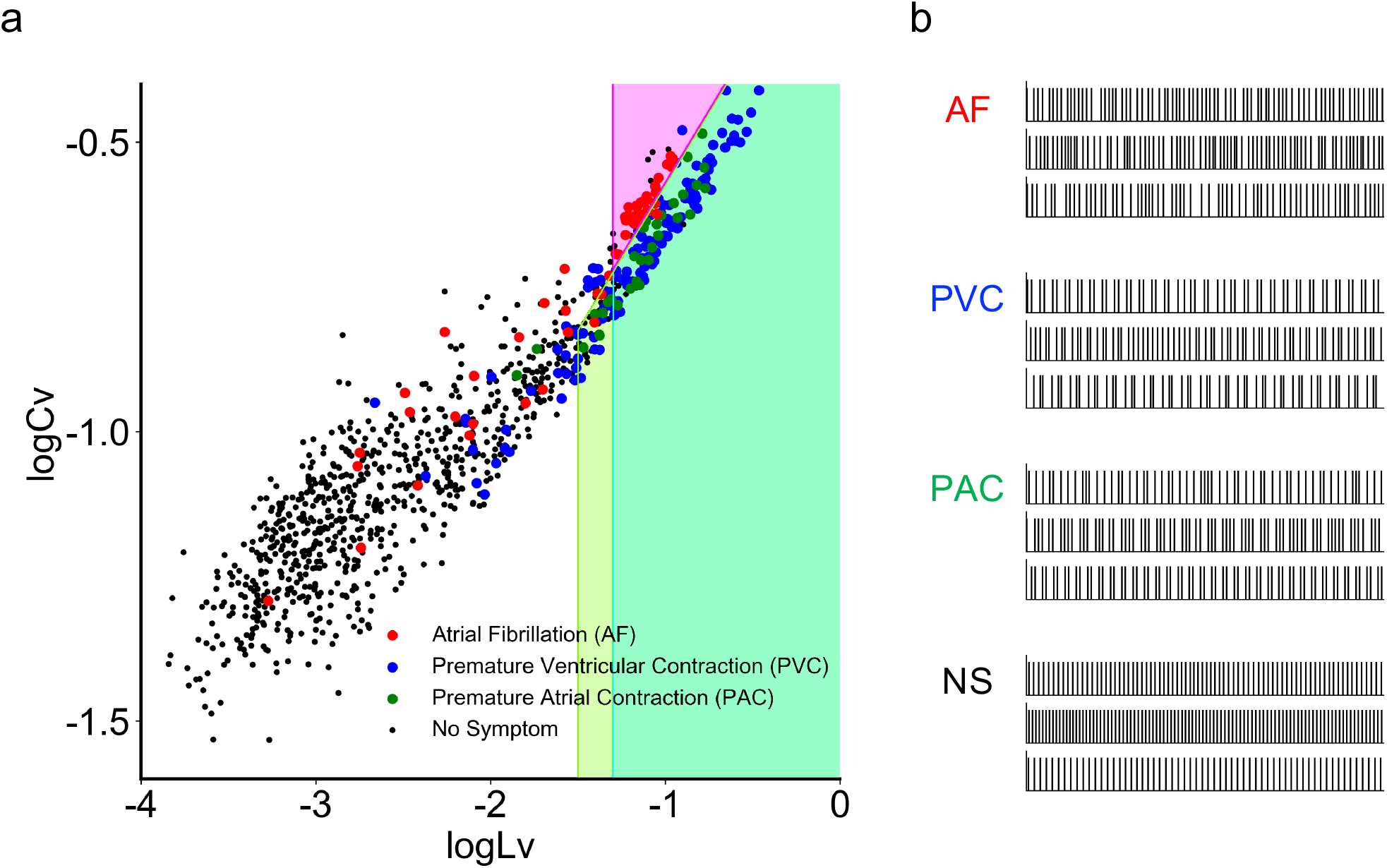
Different cardiac disorders plotted on a plane spanned by the new and old metrics log_10_ *Lv* and log_10_ *Cv*. (a) Dots represent 1,017 subjects, who were diagnosed as atrial fibrillation (AF), premature ventricular contraction (PVC), premature atrial contraction (PAC), and negative, respectively colored in magenta, blue, and green, and black. The prediction zone for each cardiac disorders, given by the set of constraints log_10_ *Lv > a* and log_10_ *Lg* = log_10_ *Lv* − 2 log_10_ *Cv > b* (or *< b*) is depicted by the appropriate colors. (b) Representative pulse sequences of 1 minute, respectively taken from four different cases. NS stands for no symptom.

Measurements associated with cardiac disorders are located in the upper right region defined by high values of log_10_ *Lv* and log_10_ *Cv*. On further analysis, the data points representing different cardiac disorders are distributed differently in this region; fibrillation (AF) cases are located on the upper side, whereas premature contraction (PVC and PAC) cases are located on the lower side. Representative pulse sequences of 1 minute taken from these diagnostic cases are depicted in Fig. 3(b).

To determine how the local variation *Lv* and the coefficient of (global) variation *Cv* contribute to the separation of premature contractions and fibrillation, we newly introduce a metric of the local-global variation ratio (*Lg*), defined as

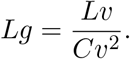

In the denominator, *Cv*, which measures the standard deviation of intervals, is squared to conform to the power of deviation in the numerator *Lv* measures the squared deviation (METHODS). For a Poisson random pulse train, *Lg* exhibits the value unity because both *Lv* and *Cv* take the value unity. For a locally regular pulsation whose rate is slowly modulated, *Lg* ≪ 1, because *Lv* is much smaller than *Cv*^2^. For a perfectly regular pulsation in which RR intervals are identical, *Lg* is undefined because both the numerator *Lv* and the denominator *Cv*^2^ are zero. We shall prove in the METHODS that *Lg* takes the value 3*/*2 for a sequence in which RR intervals of similar durations are arranged randomly. For a pulsation in which long and short intervals alternate, however, *Lg* takes the even higher value 3. In this way, the local-global variation ratio *Lg* can discriminate how intervals are arranged even if the intervals are close to each other (as is the case with heartbeats).

It is noteworthy that in Fig. 3, premature contractions (PVC and PAC) and fibrillation (AF) are well separated by the line log_10_ *Lg* = log_10_ *Lv* − 2 log_10_ *Cv* ≈ 0.15. It is helpful to recall that *Lg* = 3*/*2 for randomly arranged RR intervals and that log_10_ 3*/*2 ≈ 0.18. As we have seen, *Lg* can be as large as 3 if long and short intervals alternate. This means that long and short pulse intervals tend to alternate in premature contractions, as they are distributed on the lower side of the line log_10_ *Lg* = log_10_ *Lv* − 2 log_10_ *Cv* ≈ 0.18.

Considering these features, it is advantageous to infer the presence of each cardiac disorder based on a combination of variability metrics given by the following set of constraints,

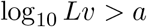

and

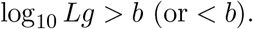

We selected the parameters *a* and *b* and the inequality direction so that the goodness index MCC is maximized for each cardiac disorder. The selected parameters are as follows: (PVC): log_10_ *Lv >* −1.3, log_10_ *Lg >* 0.14; (PAC): log_10_ *Lv >* −1.5, log_10_ *Lg >* 0.15; (AF): log_10_ *Lv >* −1.3, log_10_ *Lg <* 0.15. The selected zones are depicted in different colors in Fig. 3.

The method of combining *Lv* and *Lg* achieves MCC performances much higher than those obtained from simple thresholding of either *Cv* or *Lv* alone (Table I). In particular, the ability to detect fibrillation (AF) was drastically improved by the combination method. Note that the obtained MCC performances would practically be unchanged even if we make leave-one-out cross-validation because the number of parameters (one or two) is much lower than the number of data points (higher than 1,000).

**TABLE I.** MCC values measuring the performances of each detection method.

| Detection method | PVC | PAC | AF |
| --- | --- | --- | --- |
| $Cv$ | 0.54 | 0.30 | 0.33 |
| $Lv$ | 0.55 | 0.32 | 0.26 |
| Combination | 0.63 | 0.38 | 0.59 |

### C. Detecting cardiac disorders based on a shorter recording period

So far we have compared detection methods using 18-hour data of RR intervals extracted from 1-day Holter ECG. As the Holter recording is costly and burdensome, it would be ideal to be able to detect cardiac disorders even with a shorter interval. Here we are interested in how the detection performance degrades if the recording duration is shortened, such as 1 hour, 10 minutes, or even 1 minute. We clipped out such shorter recordings from full-length data and estimated the MCC values for detection performance using the single metrics *Cv* and *Lv* as well as the combination method. Figure 4 shows that our new combination method may provide good performance even based on shorter recordings, with an accuracy comparable to or even higher than that obtained by applying a conventional single metric such as *Cv* to an 18-hour data of RR intervals.

**FIG. 4.**
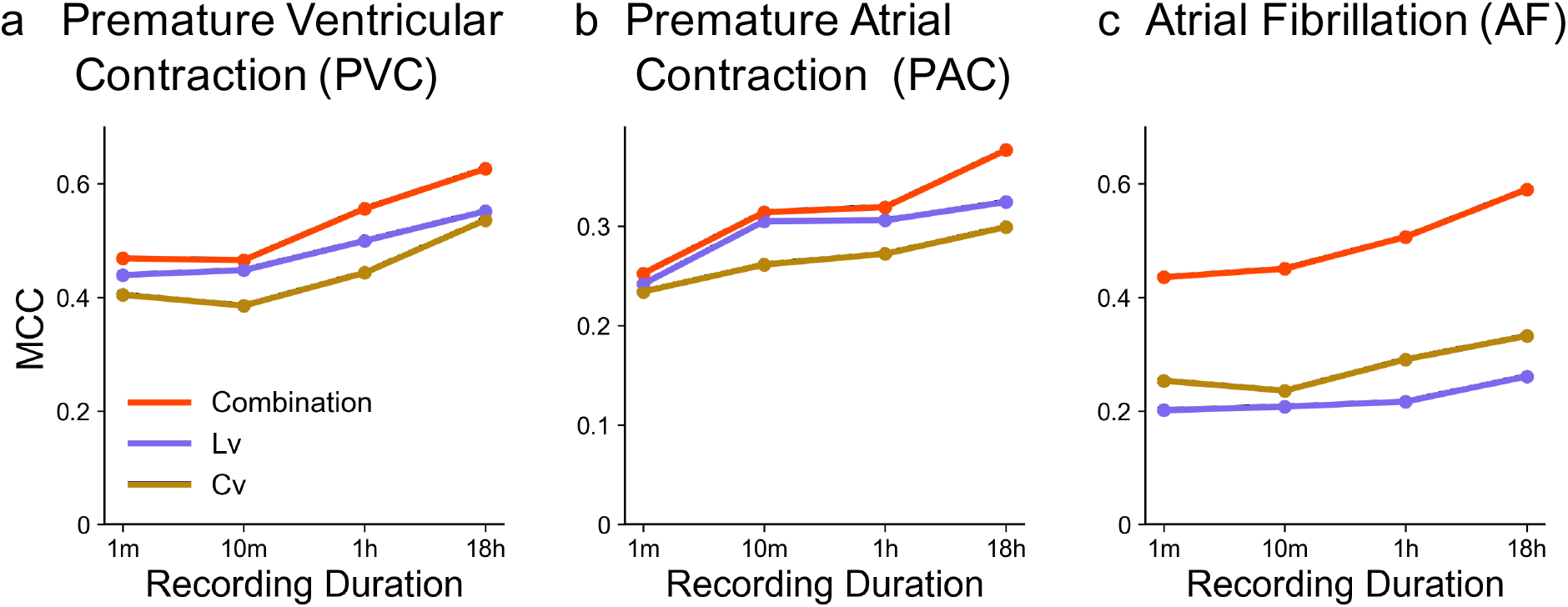
The MCCs for *Cv* and *Lv*, and the combination method based on datasets with various recording durations: 1 minute, 10 minutes, 1 hour, and 18 hours. (a) PVC, (b) PAC, and (c) AF.

Figure 5 depicts distributions of HRV metrics computed from recording periods of 1 minute, 10 minutes, and 1 hour. While datasets of shorter durations are more scattered on a plane spanned by log_10_ *Lv* and log_10_ *Cv*, the distributions of premature contraction (PVC and PAC), fibrillation (AF), and negative cases remain well separated.

**FIG. 5.**
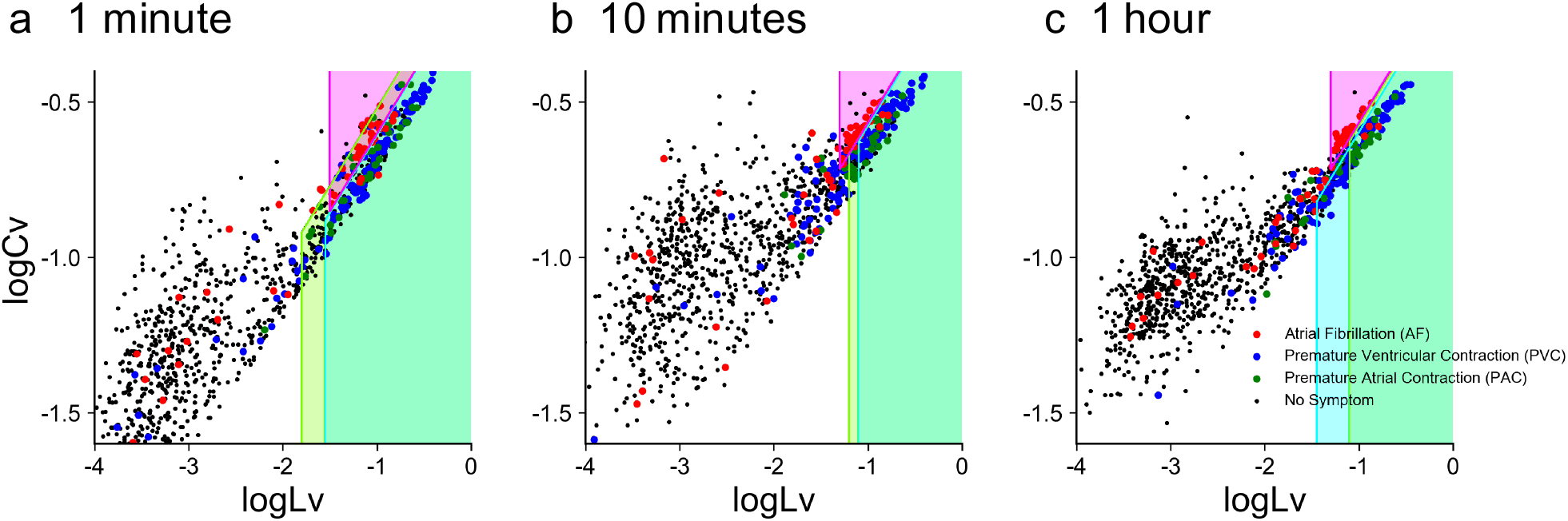
Two metrics log_10_ *Lv* and log_10_ *Cv* computed from shorter recording periods. (a), (b), and (c): Datasets of 1 minute, 10 minutes, and 1 hour, plotted on a plane spanned by log_10_ *Lv* and log_10_ *Cv*. The parameters *a* and *b* of each prediction zone were adapted to respective datasets.

### D. Robustness against erroneous observation

If we monitor pulsations using a handy device such as a smartwatch, the measurement will be inaccurate since the pulse intervals are not identical to RR intervals. Considering possible fluctuations in the measurements, we created noisy data by jittering the original RR intervals with noises of the mean zero and the standard deviations of 10, 50, 100, 200, and 500 ms. Figure 6 demonstrates the manner in which the MCC values degrade with different noise durations. We can see that the combination method withstands the fluctuations if their noise level is less than 100 ms.

**FIG. 6.**
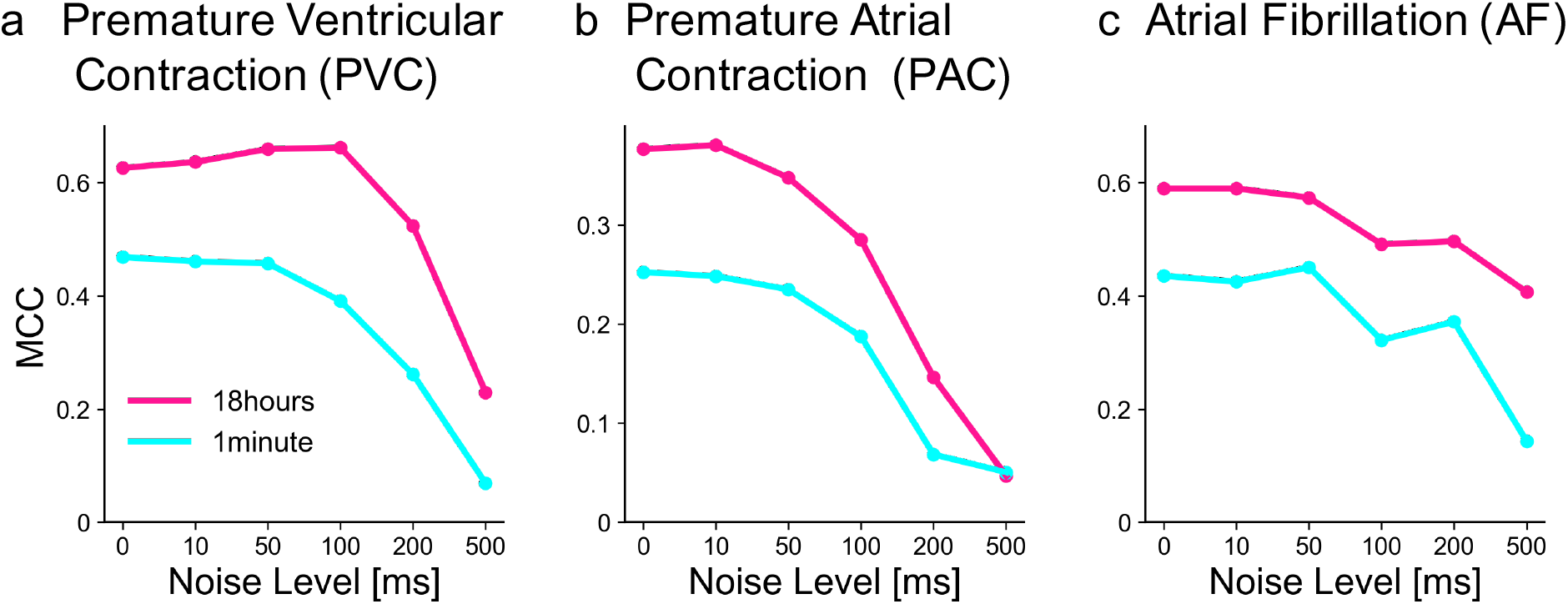
The MCCs for the combination method applied to recording duration of 1 minute and 18 hours. (a) PVC, (b) PAC, and (c) AF. Original pulse times were jittered with Gaussian noises of the standard deviation of 10, 50, 100, 200, and 500 ms.

Figure 7 depicts HRV metrics of noisy datasets, in which the original series of RR intervals of 18 hours were jittered with noises of the standard deviations of 10, 50, and 100 ms. While datasets of noisy RR intervals were shifted toward higher values of log_10_ *Cv* and log_10_ *Lv*, the distributions of premature contraction (PVC and PAC), fibrillation (AF), and negative cases remain well separated for these cases.

**FIG. 7.**
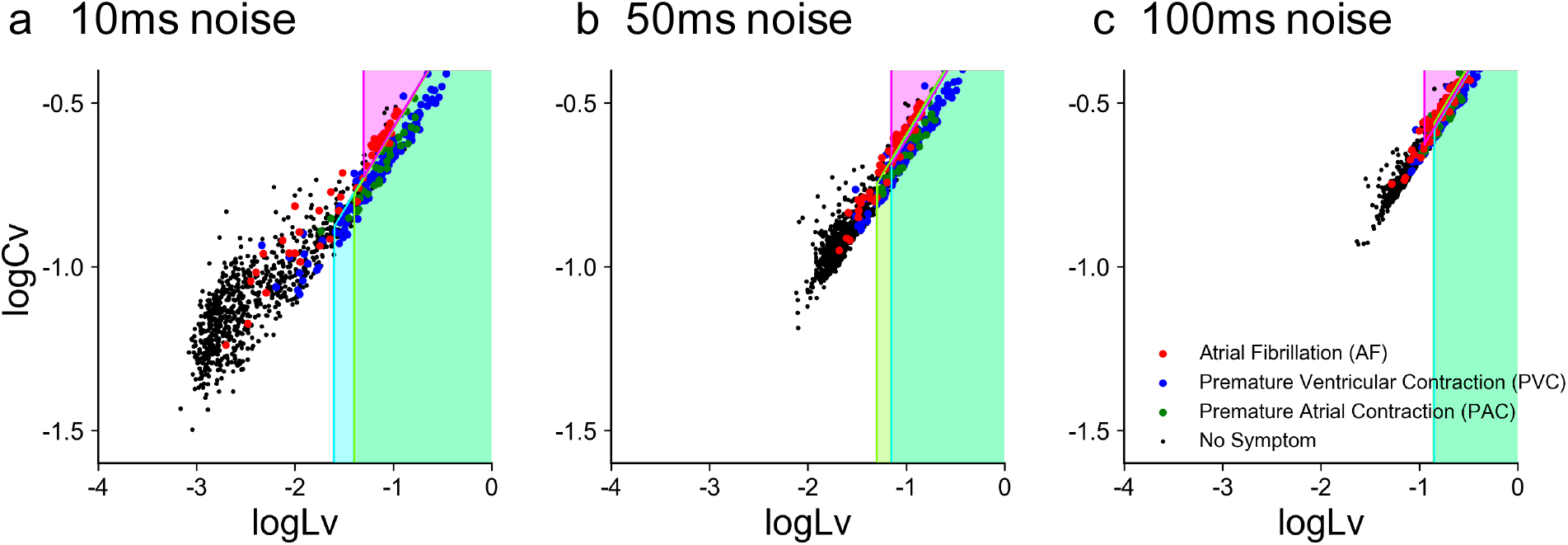
Noisy datasets of different cardiac disorders plotted on a plane spanned by log_10_ *Lv* and log_10_ *Cv*. (a), (b), and (c): Datasets were obtained by jittering original pulse times with Gaussian noises of the standard deviation of 10 ms, 50 ms, and 100 ms. The parameters *a* and *b* of each prediction zone were adapted to respective noisy datasets.

### E. Application to public databases

To examine if our method may also be valid for other datasets, we applied it to the data of the MIT-BIH databases, which have been deemed standard. Here we considered the “Atrial Fibrillation Database” as representing the symptoms similar to the AF cases selected by Kyoto Industrial Health Association, while the “Arrhythmia Database” and the “Normal Sinus Rhythm Database” represent those who were not AF. Figure 8 represents the distributions of log_10_ *Lv* versus log_10_ *Cv* of 1minute, 10minute, 1hour, and 18hour data of the MIT-BIH datasets. For each dataset, two parameters *a* and *b* of our prediction model were selected so that MCC is maximized. The MCC values obtained for the AF cases of MIT-BIH datasets were much higher than those obtained for our Kyoto datasets; for instance, the MCC value for 18-hour data was as high as 0.761. The difference may have arisen because diagnostic criteria and the population ratio of cardiac cases were different between these datasets. Nevertheless, it is noteworthy to see the two parameters *a* and *b* were similar to those of Kyoto database; for instance, *a* = −0.115 and *b* = 0.27 for 18 hour data. This implies that a similar criterion may be applicable even for different datasets.

**FIG. 8.**
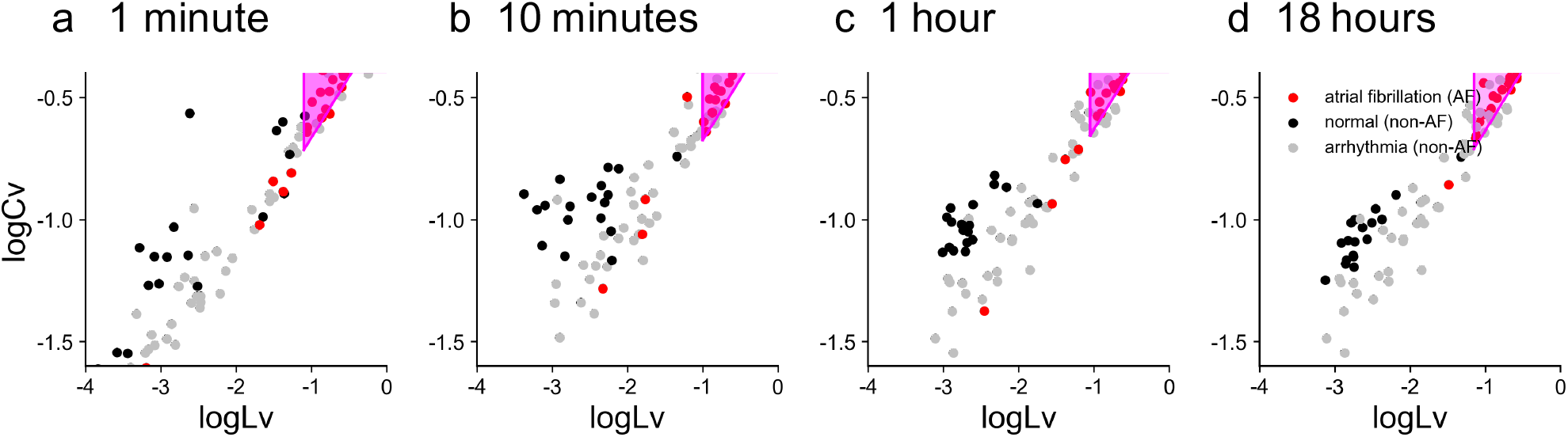
Two metrics log_10_ *Lv* and log_10_ *Cv* computed for three kinds of MIT-BIH databases. (a), (b), (c), and (d): Datasets of 1 minute, 10 minutes, 1 hour, and 18 hours plotted on a plane spanned by the two metrics. The parameters *a* and *b* of the prediction zone (magenta) for the AF cases were adapted to respective datasets.

## III. DISCUSSION

In this study, we constructed an automated algorithm for detecting cardiac symptoms from pulsation signals. We first compared *Lv* with *Cv* and found that *Lv* was superior for detecting pre-mature contractions, while *Cv* was superior for identifying fibrillation. Considering the specificity of each metric, we introduced the local-global variation ratio *Lg* = *Lv/Cv*^2^ and found that this metric effectively discriminates between premature contractions and fibrillation. With a method for combining *Lv* and *Lg*, we have obtained a performance far superior to that obtained by simple thresholding with single metrics.

Considering the cost and time needed to perform 1-day Holter ECG, it is desirable to be able to make a reasonable inference from a shorter recording, such as the few-minute ECG carried out during a regular medical checkup. We confirmed that applying our new combination method to 1-minute recordings provided detection performance that was comparable to or even better than that of the conventional HRV metric *Cv* applied to a 1-day recording.

Nevertheless, this favorable performance might not be exclusive to our method. Experienced medical practitioners should be able to achieve similar performance through pulse diagnosis, or by making full use of the many existing HRV metrics. The strength of the current study might be the simplicity of the analysis and its usability; we may ease the burden of medical practitioners by making a preliminary detection of cardiac symptoms simply from the information of RR intervals.

We have seen that longer recording of pulses provides more reliable inference. This is partly because the estimation of variability metrics becomes more accurate with time. But the main reason may be that cardiac symptoms occur intermittently during 1-day recording and it is difficult to identify disorders if the analysis is performed while symptoms are absent. But if we can use a more convenient device such as a smartwatch that can constantly monitor heartbeats, cardiac disorders can be easily detected using our analysis algorithm.

By assuming that pulses reflect the RR intervals of cardiac beats, being accompanied by noises occurring downstream, we tested whether our analysis method was robust against noisy data. By jittering the original RR intervals with fluctuating noises, we have confirmed that the detection of cardiac symptoms did not deteriorate if the fluctuation in the measurements was shorter than 100 ms.

Here we selected the parameters for determining cardiac symptoms so that the performance was maximized in terms of the MCC. However, the MCC was based on a balance between true and false positives and negatives, in the sense that positives and negatives are considered to have equal weights. We may lower the threshold of *Lv* if we wish to be more cautious in predicting cardiac disorders, even though this results in an increase of false positives.

To test the generality of the analysis method, we applied it to the data of the MIT-BIH databases. We have confirmed that our method may detect AF cases with much higher MCC values, indicating the higher classification performance. It is noteworthy that we obtained a similar criterion for this dataset, although the diagnostic criteria may generally be different between different medical institutions.

The advantage of our method is its simplicity of use. Though artificial intelligence-aided methods may also achieve high performance in detecting cardiac symptoms, they have a huge number of parameters that sensitively depend on the training datasets. Our method has only two parameters *a* and *b*, and we may check whether they have been dependent on training datasets, which may be largely dependent on the diagnostic criterion of different institutions.

In this study, we achieved high detection performance by combining *Cv* and *Lv*, particularly by paying attention to the local-global variation ratio *Lg*. The essential point of improvement was to combine multiple metrics that detect different features of heartbeats. If we can obtain more sample data, it might be worthwhile to look for more detailed combinations of additional variability metrics.

## IV. METHODS

### A. ECG data

The data of Holter ECG were obtained from outpatients who had cardiological examinations at Clinic, Kyoto Industrial Health Association, Kyoto 604-8472, Japan. Japanese institutions or corporations are requested to let workers undergo a medical checkup once a year. If potentially concerning results are observed during the ECG recording performed during the physical checkup, individuals are recommended to undergo 1-day Holter ECG monitoring. Accordingly, subjects who received Holter ECG may have been more likely to have cardiac disorders than the overall population. Nevertheless, the majority of these individuals were still “healthy” in that they were not diagnosed with heart failure, and some were asymptomatic.

The ECG data obtained from a CM5 lead were analyzed. Categorization of cardiac symptoms of outpatients was performed by medical doctors who acquired the license as Physician and Surgeon in Japan. The present study was approved by Institutional Research Board, Kyoto Industrial Health Association (Permission No. S18-0006), and the Ethics Review Committee for Medical and Health Research involving Human Subjects, Ritsumeikan University. In the analysis of 1-day Holter recording, each pulse was automatically diagnosed as PVC and/or PAC by software provided by Fukuda Denshi Co. Ltd. Based on the summary data, we calculated the fraction of pulses exhibiting PVC and/or PAC contained in an entire set of pulses. Medical doctors analyzed the full records of individual subjects, and if abnormalities were detected, they manually determined the periods of AF. We calculated the ratio of the AF period to the total measurement time, 18 hours.

From the recorded RR intervals, we computed *Cv* and *Lv*. We discarded 14 datasets whose recording period was shorter than 18 hours. Accordingly, we had datasets of 1,017 subjects in total (863 independent persons, men: 625, women: 238), and analyzed the initial 18 hours. The ages of the subjects ranged from 20 to 90.

### B. MIT-BIH ECG databases

In addition to the Kyoto databases, we examined public databases MIT-BIH provided by the Harvard-MIT Division of Health Sciences and Technology [38–40]. Here we adopted three kinds of databases: MIT-BIH Atrial Fibrillation Database (2-channel Holter ECG recorded at a sampling frequency of 250 Hz for 10 hours); MIT-BIH Arrhythmia Database (2-channel Holter ECG recorded at a sampling frequency of 360 Hz for 24 hours. We adopted the 30-minute waveform data); and MIT-BIH Normal Sinus Rhythm Database (2-channel ambulatory ECG recorded at a sampling frequency of 128 Hz for 24 hours). We obtained RR intervals from the waveform data of the first channel. R-peaks were detected using the ecg peaks function of neurokit2 software [41]. For datasets whose recording period was less than the specified period (1minute, 10minutes, 1hour, 18hours), we analyzed the entire recording period.

### C. Metrics for measuring heartbeats

Given a sequence of RR intervals, we computed variability metrics defined as follows.

- Average heart rate (*Hr*) The most basic metric for measuring heartbeats is the average heart rate *Hr*, defined as,

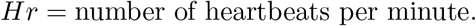 The 18-hour average value was adopted as an HR statistic for each subject.
- Coefficient of variation (*Cv*) There are many conventional approaches for detecting HRV, such as linear, frequency domain, wavelet domain, and nonlinear methods. As a representative HRV metric, we used the coefficient of variation of RR intervals, defined by

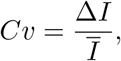

where Δ*I* and *Ī* represent the standard deviation and the mean of RR intervals. These statistics are typically measured every 10 minutes (references). Unless specified in the main text, the 18-hour mean of the logarithm of the 10-minute mean Cv was adopted as the Cv statistic. In Fig. 4, the 1-minute Cv and the 10-minute Cv were used only one-shot data by averaged over the first 1 minute and 10 minutes duration after 10-minute transient time, respectively. The 1-hour Cv was estimated as the logarithm of the 10-minute mean Cv averaged over the first 1 hour after the 10-minute transient. The coefficient *Cv* is designed to exhibit a value of unity for a Poisson random pulse train and is zero for a perfectly regular pulsation signal.
- Local variation (*Lv*) The conventional method of measuring HRV is adversely affected by slow variations in the heart rate and is also sensitive to artifacts and errors. There have been efforts to remove these artifacts, and modern methods employ machine learning techniques such as the state-space method [42, 43]. Here we suggest a simpler metric called the local variation *Lv*, which was introduced for measuring the firing irregularity of the neurons in the brain [26–28]. The local variation *Lv* is defined as,

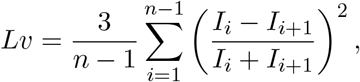

where *I*_*i*_ and *I*_*i*+1_ are the *i*th and *i* +1st RR intervals, respectively, and *n* is the total number of the intervals in a given duration. Note that the heartbeat that makes up the end of the *i*th RR interval and the start of the *i* + 1st RR interval was within the duration. The short-time measurement Lv statistics were measured in the same way as the Cv statistics. The coefficient 3 in the definition of *Lv* was chosen so that *Lv* gives the value of unity for a Poisson pulse train [26]. *Lv* is zero for a regular pulsation.

Whereas *Cv* represents the global variability of an entire sequence and is sensitive to rate fluctuations, *Lv* detects the instantaneous variability of intervals. To demonstrate the difference in the workings of these metrics, we created synthetic pulse sequences. In Fig. 9(a), RR intervals are lined up in a regular manner from long to short and then short to long, while in Fig. 9(b), the identical set of RR intervals is presented in a random sequence. Accordingly, (a) represents a locally regular pulsation while the rate is slowly modulated, whereas (b) represents irregular pulsation.

**FIG. 9.**
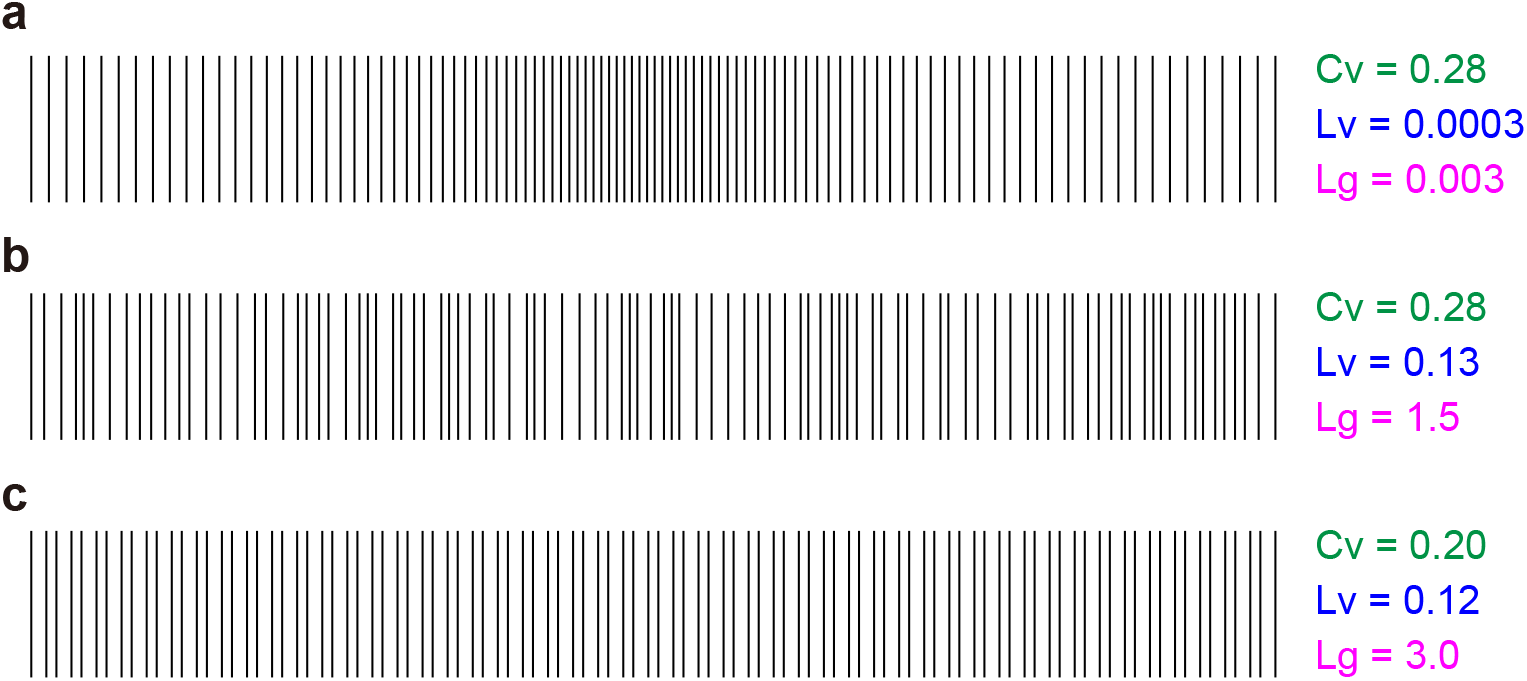
Synthetic pulse sequences and the computed values of the coefficient of variation *Cv*, the local variation *Lv*, and the local-global variation ratio *Lg*. (a): Pulsation is locally regular while the rate is slowly modulated from low to high and then back to low. (b): Pulsation is irregular, although the average rate is nearly constant. (c): Long and short RR intervals alternate with each other. *Cv* has the same value for (a) and (b), which are composed of identical sets of RR intervals, whereas *Lv* is small for (a), in which the RR intervals are well organized, and is large for (b), in which RR intervals are arranged randomly. *Lg* is very small for (a), in which the rate is slowly modulated. *Lg* ≈ 3*/*2 for (b), in which RR intervals are randomly arranged. *Lg* ≈ 3 for (c), in which long and short RR intervals alternate.

The coefficient of variation *Cv* has identical values for (a) and (b) because the standard deviation and the mean are the same for both sequences. However, *Lv* successfully ignores slow heart rate modulation, and accordingly, it identified the difference in local irregularity between the two sequences. Note that we made the variations in RR intervals much larger than those of real cardiac beats so that their differences would be apparent.

In addition to these metrics, we have newly introduced the local-global variation ratio *Lg* = *Lv/Cv*^2^. Its characteristics are analyzed in the following subsection. *Lg* takes a value much smaller than unity for a locally regular pulsation whose rate is slowly modulated (Fig. 9(a)). *Lg* takes the value 3*/*2 for a sequence in which RR intervals of mutually similar values are arranged randomly (Fig. 9(b)). *Lg* takes an even higher value of 3 for a pulsation in which long and short intervals alternate (Fig. 9(c)).

### D. Analytical calculations of *Cv, Lv*, and *Lg* for some limiting cases

#### Poisson process

For the Poisson process, in which pulses are generated at random at a given fixed-rate, pulse intervals *I*s are distributed exponentially, *p*(*I*) = 1*/τ* exp(−*I/τ*). In this case, the mean and standard deviation of pulse intervals are both *τ* and accordingly the expected value of *Cv* is

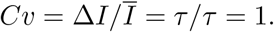

In the Poisson process, consecutive intervals appearing in the summand of *Lv* are derived independently from the same exponential distribution, and accordingly, the expected value of *Lv* is

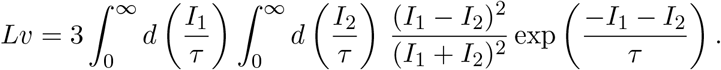

This integration can be carried out with the transformation from {*I*_1_, *I*_2_} to {*x, y*} = {(*I*_1_ + *I*_2_)/2*τ*, (*I*_1_ − *I*_2_)/*τ*} as

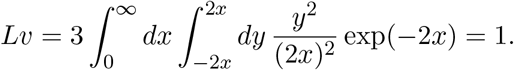

Accordingly, the local-global variation ratio *Lg* is

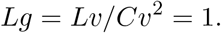

#### Renewal process with gamma-distributed interval

Expectation values are analytically available for a wide class of renewal processes in which pulse intervals are derived from the gamma distribution,

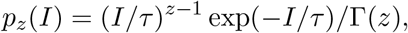

where Γ(*z*) is the gamma function defined as 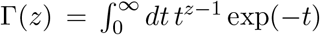. The original Poisson process corresponds to the case of *z* = 1, and we can generate more regular pulse trains with the larger *z*.

The expectation value of *Lv* is also obtained analytically as

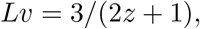

and the expectation value of *Cv* is obtained as

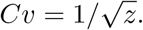

Accordingly, the local-global variation ratio *Lg* is

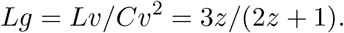

While *Lg* = 1 for the Poisson process with *z* = 1, *Lg* = 3*/*2 for a regular pulse train at a limit of *z* → ∞.

#### Sequences consisting of long and short pulse intervals

The local-global variation ratio *Lg* is undefined for a perfectly regular pulse train, for which both *Lv* and *Cv*^2^ are zero. Though we have obtained *Lg* = 3*/*2 for a regular pulse train in a limiting case of the renewal process with gamma interval distribution *z* → ∞, it might be sensitive to the arrangement of pulse intervals. Here we consider long pulse trains in which the equal numbers of long and short intervals are arranged in various orders. Let the long and short intervals be denoted as *τ* + *δ* and *τ* − *δ*, respectively (*τ > δ >* 0). Because the mean interval and standard deviation are *τ* and *δ*, respectively, the coefficient of variation *Cv* is

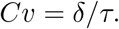

By denoting the switching probability between long and short intervals as *p*, the local variation *Lv* is given

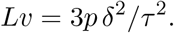

Accordingly, the local-global variation ratio *Lg* is obtained as

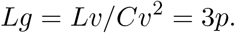

Thus *Lg* = 3*/*2 if long and short intervals alternate randomly, in which case the switching probability between long and short is half, *p* = 1*/*2. But *Lg* can be as high as 3 if the long and short intervals always alternate (*p* = 1). *Lg* can be close to zero for a long pulse train in which switching between long and short intervals occurs very rarely (*p* ≈ 0). In this way, the local-global variation ratio may discriminate the order of long and short intervals, even if the difference between the long and short intervals is very small (*δ* ≪ *τ*).

## Data Availability

All data produced in the present work are contained in the manuscript

## ACKNOWLEDGMENTS

We thank Luis Rocha, Hiroshi Matsuura, and Masahiro Naito for their support at the preliminary phase of this analysis. We are particularly grateful to Noboru Imagawa for preparing data at Kyoto Industrial Health Association.

